# Potential limitations of community-wide strategy to treat Mycobacterium tuberculosis infection

**DOI:** 10.64898/2026.05.13.26353093

**Authors:** Saikat Batabyal, Kevin Urdahl, Vitaly V. Ganusov

## Abstract

A quarter of the world’s population has immunologic evidence of past or present Mycobacterium tuberculosis (**Mtb**) infection (**MTBI**) detected as TST or IGRA positivity. Community-based preventive treatment of individuals with MTBI has resulted in transient decreases in TB cases, but its long-term effectiveness has been controversial. Due to the likelihood that many of those with immune responses to Mtb antigens may no longer harbor Mtb, widespread treatment of all such individuals may result in unnecessary exposure to antibiotics. We raise an additional concern that preventive treatment of individuals with MTBI, who are not at the risk of disease progression, may result in loss of protective immunity, provided by the persistent infection, and enhanced risk of TB upon re-exposure to Mtb. There is evidence from human cohorts and animal studies that prior exposure to Mtb confers protection against TB development upon re-exposure, and that treatment of Mtb-infected animals often results in loss of this protection. We build a novel epidemiological model of Mtb dynamics and progression to TB in a community allowing for protection afforded by MTBI against exogenous reinfection-driven disease progression. We show that implementation of treatment of MTBI in the whole community will result in reduction of TB cases but stopping the program may result in an increase in new TB cases that may offset (or even exceed) benefits of the preventive treatment program. Our results suggest that better understanding protective effects provided by MTBI against progression to TB upon Mtb re-exposure and identification of Mtb-infected individuals who most benefit from preventive treatment must be a priority before preventive treatment of asymptomatic MTBI is widely implemented.

## Introduction

Tuberculosis (**TB**), a disease caused by Mycobacterium tuberculosis (**Mtb**), remains a major global health concern with more than 10 million cases of TB and over 1 million deaths due to TB annually^1^. In the absence of an effective vaccine, the major strategy of controlling TB incidence has been by identifying and treating individuals with active disease^1–5^. This approach has not been successful in developing countries where TB incidence has been relatively stable in the last two decades suggesting that additional approaches are needed if we are to sharply reduce TB incidence in the world^1^.

Preventing TB development in individuals with evidence of exposure to Mtb is an attractive additional strategy to reduce TB incidence^6–9^. Prevalence of Mtb infection (**MTBI**), detected as tuberculin skin test (**TST**) or interferon gamma release assay (**IGRA**) positivity, varies in different regions and countries and typically increases with age, reaching 70-80% in some communities^10–17^. Multiple studies have established that preventive (or prophylactic) treatment of MTBI^+^ contacts of TB index cases is a highly effective strategy to prevent TB development in the contacts^18,19^. Yet, identifying individuals with a recent exposure to Mtb is difficult and expensive especially in high TB burden settings^20,21^.

An alternative strategy to control TB incidence in a population is TB preventive treatment (**TPT**) of all MTBI^+^ individuals^6,8,9,22^. Indeed, a mathematical modeling-based analysis suggested that combining TPT and treatment of individuals with active disease would act synergistically to reduce TB incidence^6^. The key assumption of this (and many other) mathematical models is that MTBI puts an individual on a path of progression to active disease, driven primarily by endogenous reactivation^23–25^. In such models preventive treatment stops disease progression by eliminating Mtb^26^. However, if disease progression is determined primarily by new exposure to Mtb (i.e., exogenous reinfection), the benefits of community-wide preventive treatment of MTBI^+^ individuals may be overestimated.

The relative importance of endogenous reactivation or exogenous reinfection in driving TB incidence in a population has been debated for decades^27,28^. A relatively rapid decline in TB incidence among US immigrants from TB endemic countries suggested importance of exogenous reinfection in the country of origin^29^; some studies by using Mtb genotyping suggested that exogenous reinfection contributes from 40 to 90% of new TB cases in USA, Spain, or South Africa^30–32^. In contrast, in China (Hong Kong) most of new TB cases in the elderly (75%) have been attributed to endogenous reactivation^33^.

Experimental evidence of community-wide benefits of preventive treatment of individuals with MTBI is mixed^34–36^. A large collection of randomized clinical trials tested benefits of year-long TPT with isoniazid (**INH**) on TB incidence in settings of low or high Mtb transmission (e.g., contacts of TB cases, individuals in US mental institutions, or Alaska villagers); INH treatment reduced the overall TB incidence in 5-10 years by about 50% in most studies^34^. The major impact of the preventive treatment was in the first year of the trials, when INH was administered, and yet INH-treated cohorts generally experienced fewer cumulative TB events as compared to placebo-treated groups even when excluding first few years after treatment from the analysis^34^. In more recent trials in settings of high Mtb exposure preventive treatment reduced a chance of progression to active disease^35,36^; however, stopping the treatment typically resulted in resurgence of TB cases in previously treated individuals. Overall, given that the long-term benefits of the community-wide treatment of MTBI have not been demonstrated, current guidelines in several countries do not recommend community-wide preventive treatment^37–39^.

Not all individuals with MTBI may benefit from preventive treatment. MTBI^+^ individuals likely represent a highly heterogeneous group: some individuals with MTBI may no longer harbor the bacteria but have immune memory from the past infection, others may harbor the bacteria along with associated immunity, and others may be on the way of progression to active disease^5,40,41^. The risk of TB progression is highest in the first two years following primary exposure to Mtb^42^; however, only a small proportion of Mtb-exposed individuals will progress to active disease and thus would benefit from preventive treatment. In particular, *<* 2% of individuals in the Prophit survey in the UK progressed to active disease in a 10 year follow-up as compared to 14.5% of Mtb-exposed individuals in more recent study in Australia^43,44^. Importantly, those individuals who did not progress to active disease upon Mtb exposure may derive benefits if re-exposed to Mtb (i.e., due to exogenous reinfection). Indeed, multiple cohort studies identified reduced or delayed progression to active disease of individuals with positive TST in situation of high Mtb exposure, e.g., household contacts of TB cases, nursing students attending TB wards, or elderly in nursing homes^43,45–52^. These observations have been confirmed in animals studies. For example, contained Mtb (**CoMtb**) infection of mice or low dose/non-progressive Mtb infection of monkeys is also highly protective when infected animals are re-exposed to Mtb^53–55^. However, antibiotic treatment of the CoMtb animals results in loss of protection^54,56^, and treated MTBI^+^ individuals lose Mtb-specific immunity^57^ suggesting that preventive treatment of individuals with MTBI may in fact result in loss of MTBI-induced protection and increased susceptibility to Mtb upon re-exposure.

By using mathematical modeling we sought to investigate the potential impacts of preventive treatment of individuals with MTBI on TB incidence in a community. Mathematical modeling has been used extensively to understand TB epidemiology and to quantify impact of various interventions on TB incidence^6,23,24,58–62^. Our novel mathematical model differs from previous versions in two key ways. First, we specifically consider individuals with MTBI as those progressing to active disease and those who are not progressing but derive benefits from CoMtb upon re-exposure to Mtb^63–65^. Second, we allow individuals in the non-progressive state to heal and eventually revert to the MTBI-negative (i.e., TST^*−*^) state^66^. Recent analyses suggest that nearly all (if not all) mathematical models of TB progression do not allow “reversions” (or cure) of infected individuals to the naive/TST^*−*^ state^23–25^ despite abundant experimental evidence of loss of TST positivity in cohort studies ^25,67,68^. Our results suggest that preventive treatment of individuals with MTBI will result in reduced TB incidence in the community; however, stopping the program without substantial reduction in the force of Mtb infection will result in increased TB incidence that may significantly exceed values observed prior to the treatment program start. Our results thus suggest that benefits of implementing preventive treatment programs in large communities must be carefully weighted against potential increases in TB incidence due to loss of protection of treated individuals with controlled, non-progressing, and protective MTBI.

## Materials and methods

### Data

The data on progression of nursing students to TB have been published previously^47,48^; we entered the data provided in Table 4 of Badger & Ayvazian ^48^ into csv format (available on GitHub). In short, Badger & Ayrazian ^47^ followed 745 students in a nursing school in Boston, MA (USA) over the period of 15 years. The first three years students spent at the school, and remaining time they were working in other parts of the US. Upon entry to the school, 362 students were TST^*−*^ and 374 were TST^+^ with TST reactivity being unknown for 9 students. TST^+^ students with evidence of TB were not included in the cohort. Out of 362 TST^*−*^ students 285 developed TST positivity (and 52, or 14%, did not); the majority of the 40 TST^*−*^ students that did progress to active disease converted to TST positivity within two years (**Supplemental Figure S2**A). Over the course of 15 years, 31 or 40 of TST^+^ or TST^*−*^ students, respectively, developed TB (**Supplemental Figure S2**B-C). There was no reported data on how quickly TST positivity developed in the remaining 245 students who did not develop active TB in the 15 year follow up period. We used these data to constrain parameters of the epidemiological model of Mtb dynamics (see below); a more detailed analysis of the progression to TB data will be presented elsewhere.

### Mathematical models

#### Mathematical model to describe progression to TB and protection by MTBI

In the model we assume that susceptible individuals (*S*) upon exposure to Mtb (at a rate *βT*) either become latently infected *L* (and thus are non-progressing to disease) or enter a progressive state *I* (and are on the path towards active disease). The proportion of susceptible individuals entering the progressive state per exposure is *η≪* 1. Individuals in the progressive state progress to active disease at rate *θ* + *βT* that combines the effect of progression following primary exposure (*θ*) and due to re-exposure to Mtb (*βT*). Individuals in the non-progressive convert into the progressive state upon re-exposure to Mtb at a rate (1*− ϵ*)*βT* where *ϵ* is the degree of protection afforded by MTBI; *ϵ* = 1 when MTBI induces a fully protective state. Individuals in the progressive state can spontaneously control the infection and revert into non-progressive state at rate *λ*. Implementation of the community-wide program of treating MTBI would move individuals from the progressive and non-progressive states into susceptible state at a rate *δ*_*L*_. The mathematical model is written as follows:

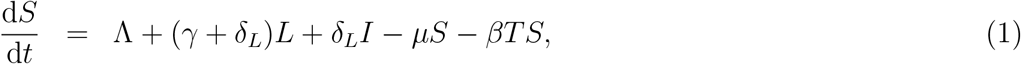

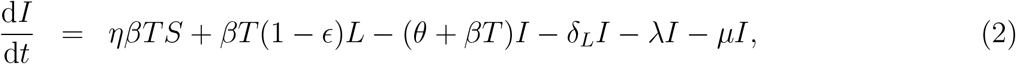

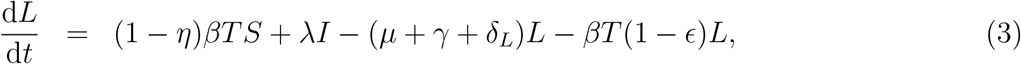

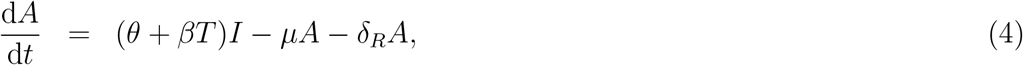

where Λ is the rate of production of new susceptible individuals (birth rate), *µ* is the background death rate, *γ* is the rate at which individuals in non-progressive state lose MTBI-induced protection and become fully susceptible to reinfection by Mtb, *δ*_*R*_ is the rate of death/removal of individuals with active disease, *β* is the infection rate and *T* is the total number of infectious individuals in the population (**Figure 1**). We assume that the community under consideration for preventive treatment is small in comparison to the total number of infectious individuals in the overall population (i.e., at the steady state *A*^*\**^ *≪ T*), so the force of infection *βT* is not strongly impacted by the reduction in active cases during treatment^62^. We chose parameters so the total population size at the steady state prior to treatment is close to 1 (*S*^*\**^ + *I*^*\**^ + *L*^*\**^ + *A*^*\**^ *≈* 1). In model simulations, we assume that preventive treatment of asymptomatic individuals (*I* + *L*) as the population is at a steady state, i.e. we initially let *δ*_*L*_ = 0. The equilibrium state of the population prior to treatment program start is as follows:

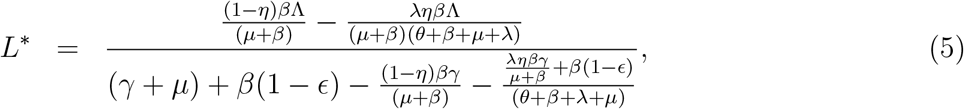

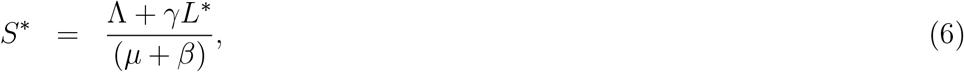

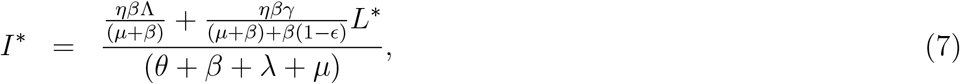

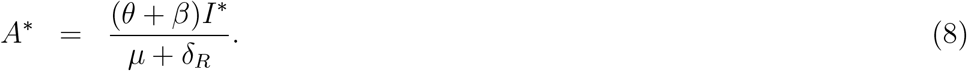

**Figure 1:**
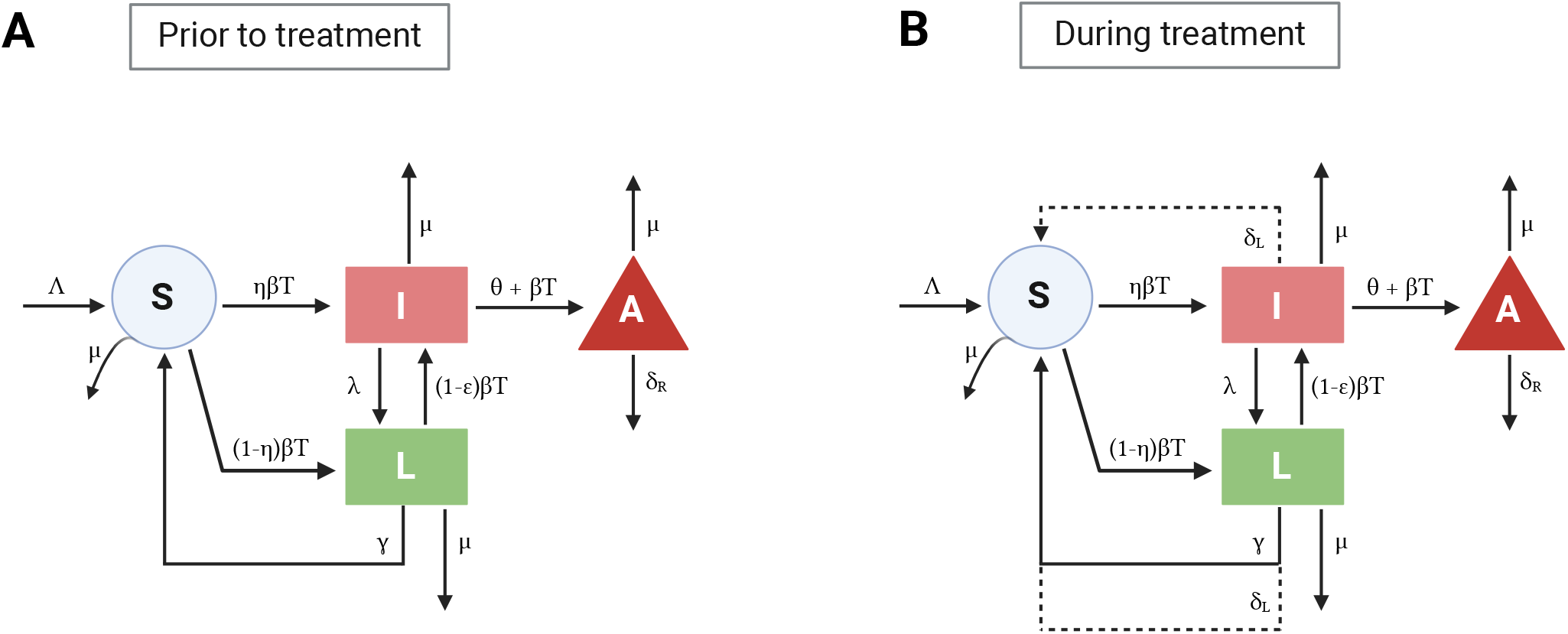
Schematic diagram of a simplified mathematical model of progression to TB with MTBI-induced protection. In the model (**eqns. (1)–(4)**) we follow the relative numbers of individuals at different stage of the disease prior (**A**) or during the treatment (**B**). There is a constant birth rate of susceptible individuals (at rate Λ), and all individuals have a background death rate *µ*, and individuals with active disease (*A*) are treated and/or die at a rate *δ*_*R*_. **A**: susceptible individuals *S* upon exposure to Mtb at a rate *βT* either move into progressive state *I* or non-progressive state *L* with *η* being the probability for an individual to move into progressive state (typically, *η ≪* 1). Individuals in the progressive state progress to the active disease state *A* determined by the intrinsic rate *θ* and the rate of re-exposure to Mtb *βT*. Also, individuals in the progressive state may spontaneously control the infection and revert to non-progressive state at rate *λ*. Individuals in the non-progressive state may also transition into progressive state following re-exposure to Mtb at a rate (1*− ϵ*)*βT* where *ϵ* is the degree of protection of individuals afforded by the MTBI. Individuals in a non-progressive state also lose their protective status and become fully susceptible to reinfection with Mtb at a rate *γ*. **B**: Implementation of the preventive treatment program of MTBI moves individuals from both progressive and non-progressive states to susceptible state at a rate *δ*_*L*_ (shown by dashed lines). We also considered an alternative model that allows for *n* stages (*n ≥* 1) in the progressive and non-progressive states (**Supplemental Figure S1**).

Furthermore, we obtain the following characteristic equation of **eqns. (1)–(4)**:

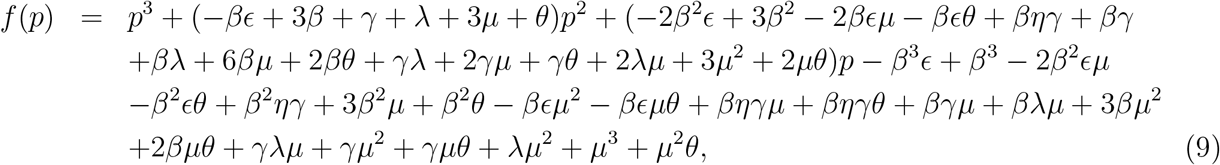

where *p*^*′*^*s* are the associated eigenvalues found by solving *f* (*p*) = 0. Finding eigenvalues and their corresponding eigenvectors analytically was very lengthy and the solutions did not fit within a page and thus are omitted here. We introduce preventive treatment of asymptomatic individuals at time 0 (with a rate *δ*_*L*_) and assume that the treatment policy is implemented for some time period (e.g., 10 years). We follow changes in the relative number of individuals in different disease stages prior, during, and after treatment policy.

#### Model parameters

Because our model is supposed to represent a generic community several of the model parameters were chosen relatively arbitrary. In particular, we assume natural birth (Λ) and death (*µ*) rates to be such that in the absence of disease (*T* = 0), the total population size at the steady state is one. Assuming life expectancy of uninfected individuals of 75 years results in the natural death rate *µ* = 0.013*/*year and Λ = 0.013*/*year (**Table 1**). The likelihood of progressing to active disease upon exposure to Mtb (*η*) depends on various factors such as intensity of exposure, age, and immune status of an individual^43,69–72^; in our simulations we let *η* = 0.05 as it has been generally stated^71^. It has been relatively well established that those individuals who progress relatively rapidly to active disease, do so within 1-2 years after the exposure to Mtb^66,73–75^. We therefore adjusted the parameter *θ* to a value that matches the rate of progression of nursing students to TB in our dataset; value *θ* = 2*/*year accurately described our data for both TST^*−*^(*ϵ* = 0) and TST^+^ (*ϵ >* 0) individuals (**Supplemental Figure S2**). The rate at which individuals in a progressive state self-control the infection (*λ*) is not well understood but may be substantial under some conditions (e.g., rest and proper nutrition)^43,76,77^, we let *λ* = 1*/*year. Natural mortality due to TB depends strongly on multiple factors such as age, immune and nutritional status; in our simulations it was set to *δ*_*R*_ = 0.3*/*year^78,79^.

**Table 1:** Parameters of the basic mathematical model used to simulate Mtb dynamics in humans population and impact of preventive treatment of asymptomatic individuals. We list parameters used in simulations (see **eqns. (1)–(4)**) as high transmission/high recovery and low transmission/low recovery; both sets are primarily determined by parameters *βT* (transmission rate) and *γ* (the rate of loss of MTBI-induced protection of individuals in non-progressive state). Other parameters include birth rate Λ, background death rate *µ*, the probability of an Mtb-exposed individual to progress to active disease *η*, transmission rate *βT* with *T* being a total number of infectious individuals in the population, the rate of loss of protection of individuals with MTBI *γ*, the rate of progression of infected individuals towards active disease *θ*, the rate of sponteneous control of progression *λ*, the rate of removal (death) of individuals with active disease *δ*_*R*_, the rate of preventive treatment of asymptomatic individuals *δ*_*L*_, and the degree of protection afforded by MTBI of individuals in non-progressive state *λ* (see Materials and methods for justification of the specific values of the parameters).

| Representation of parameters | Parameters | High infection rate $\beta T$<br>High recovery rate $\gamma$ | | Low infection rate $\beta T$<br>Low recovery rate $\gamma$ | |
| --- | --- | --- | --- | --- | --- |
| Birth rate | $\Lambda$, 1/y | 0.013 | 0.013 | 0.013 | 0.013 |
| Death rate | $\mu$, 1/y | 0.013 | 0.013 | 0.013 | 0.013 |
| Fraction to progressive state | $\eta$ | 0.05 | 0.05 | 0.05 | 0.05 |
| Infection rate | $\beta T$, 1/y | 0.05 | 0.05 | 0.01 | 0.01 |
| Recovery rate | $\gamma$, 1/y | 0.3 | 0.3 | 0.05 | 0.05 |
| Progression rate to disease | $\theta$, 1/y | 2 | 2 | 2 | 2 |
| Reversion to non-progressive state | $\lambda$, 1/y | 1 | 1 | 1 | 1 |
| Rate of active TB to be removed | $\delta_R$, 1/y | 0.3 | 0.3 | 0.3 | 0.3 |
| Rate of preventive treatment | $\delta_L$, 1/y | 1.5 | 1.5 | 1.5 | 1.5 |
| Infection-induced protection | $\epsilon$ | 0 | 1 | 0 | 1 |
| **% TB** | - | **$\epsilon = 0$** | **$\epsilon = 1$** | **$\epsilon = 0$** | **$\epsilon = 1$** |
| Prior to treatment (Steady states) | - | 1.74% | 0.46% | 0.35% | 0.09% |
| During treatment (Nadir) | - | 0.58% | 0.35% | 0.09% | 0.07% |
| After treatment (Peak) | - | 1.75% | 0.46% | 0.35% | 0.1% |

Preventive treatment of asymptomatic individuals with isoniazid takes 6-9 months with newer regimes involving two drugs may be shorter^80^; we assume that the treatment rate *δ*_*L*_ = 1.5*/*year. The degree of protection afforded by MTBI *ϵ* was varied between 0 (no protection) and 1 (full protection).

For the remaining model parameters we considered scenarios of high and low Mtb transmission settings (**Table 1**). In high transmission setting, TB prevalence is relatively high and is driven primarily by the high infection rate *βT* = 0.05*/*year. To make sure that TB prevalence is reasonable (e.g., few percent of the total population), we had to assume a high rate at which individuals in a non-progressive state lose protection (*γ* = 0.3*/*year). Unfortunately, the actual rate at which Mtb-infected individuals in a non-progressive state lose their protective status has not been accurately estimated and may depend on variety of factors. Cohort studies estimated that TST^+^ individuals lose their TST reactivity over time, with a rate of about 10%/year^25,43^. At these parameters the proportion of individuals with active disease is 1.75% (with *ϵ* = 0) or 0.46% (with *ϵ* = 1), and proportion of individuals with MTBI is 12.2% and 13.4% for *ϵ* = 0 or *ϵ* = 1, respectively (**Table 1**). In low transmission setting, we let *βT* = 0.01*/*year and *γ* = 0.05*/*year (**Table 1**). At these parameters the proportion of individuals with active disease is 0.35% or 0.09% for *ϵ* = 0 or *ϵ* = 1, respectively, and the proportion of individuals with MTBI is 12.2% and 13.3% for *ϵ* = 0 or *ϵ* = 1, respectively (**Table 1**).

## Results

### Novel mathematical model of TB progression accounting for protection by MTBI against exogenous reinfection

To investigate how treatment of MTBI^+^ individuals in the TPT program may impact the overall TB prevalence in a community we built a novel mathematical model of TB progression (**Figure 1** and **Supplemental Figure S1**). Our model extends many of the previous mathematical models of TB epidemiology in two major ways^6,23,24,58–62,81^. First, we specifically divide MTBI^+^ individuals into those who are progressive to active disease and those who are not (*I* and *L*, respectively, in **eqns. (1)–(4)**). Re-infection of MTBI^+^ individuals progressing to active disease (at rate *βT*) increases the progression rate, and re-infection non-progressing individuals (at rate (1 *− ϵ*)*βT*) moves them into the progressive state at a reduced rate dependent on protective immunity induced by MTBI status. Previous models have also considered a possibility of re-infection of MTBI^+^ individuals^63–65^; however, in previous models all MTBI^+^ individuals ultimately progress to active disease. In our model *ϵ* denotes the degree of protection provided by MTBI against reinfection of individuals in the non-progressive state (**eqn. (3)**); when *ϵ* = 1, individuals in the non-progressive state are fully protected against re-infection. Second, non-progressing MTBI^+^ individuals are able to recover from the infection and become fully susceptible to reinfection. Most previous models of TB epidemiology did not allow full recovery from infection^23,24^; this is in contrast with multiple cohort studies finding that in the absence of Mtb exposure, TST positivity tends to revert over time^66^. Preventive treatment of MTBI^+^ individuals stops their progression to active disease (for individuals in the progressive state) and moves all individuals into the susceptible state (**Figure 1**B and **eqn. (1)**).

In our general model we consider that individuals in the progressive stage pass via *n* “progressive” states before reaching the active disease state (**Supplemental Figure S1** and **eqns. (S.1)–(S.6)**). An extreme case of the general model is when *n* = 1, and individuals in the progressive state become diseased at a defined rate *θ* (**Figure 1** and **eqn. (2)**). Importantly, both general and simplified models can relatively well describe TB progression of TST^*−*^ and TST^+^ nursing students (**Supplemental Figure S2**). Interestingly, even the simplified model accurately described the initial delay in TB progression of TST^*−*^ individuals (**Supplemental Figure S2**), in part because progression to active disease takes 2 stages (**Figure 1**). For TST^+^ students the simplified model predicted protection against disease progression *ϵ* = 0.65 (**Supplemental Figure S2**C). We chose other model parameters to reflect biological details of progression to TB from published studies (**Table 1** and see Materials and methods for parameter choices).

### Stopping policy to treat MTBI may result in surge of TB cases

In our simulations we consider implementation of the policy to identify all MTBI^+^ individuals in a given community and provide such individuals with a prophylactic treatment. We assume that the program is 100% efficient at identifying all MTBI^+^ individuals, and all treated individuals are fully compliant with the treatment (**eqns. (1)–(4)**). The program is run for 10 years and TB incidence is followed a follow-up of 90 years after the program end. To determine how such a program would impact TB prevalence we consider scenarios with high or low Mtb transmission (and thus, TB prevalence), and whether MTBI induces full protection (*ϵ* = 1) against reinfection or when MTBI^+^ individuals in the non-progressive state are not protected against re-infection (*ϵ* = 0, **Table 1**).

We modelled high and low Mtb transmission scenarios by changes in the force of infection *βT*; to make sure that observed frequencies of individuals with MTBI and with active disease are within biologically reasonable ranges the rate of recovery from MTBI *γ* had to be adjusted appropriately (**Table 1**). In the case of low Mtb transmission we set *βT* = 0.01*/*y and *γ* = 0.05*/*y that resulted in prevalence of active disease of 0.35% (with *ϵ* = 0) or 0.09% (with *ϵ* = 1, **Table 1**). In high transmission setting we set *βT* = 0.05*/*y and *γ* = 0.3*/*y that resulted in prevalence of active disease of 1.74% (with *ϵ* = 0) or 0.46% (with *ϵ* = 1, **Table 1**).

The dynamics of the number of individuals in different sub-populations follows similar pattern for low and high transmission settings; however, some details are different (**Figure 2** and **Supplemental Figure S3**). To facilitate comparison of changes over time, we normalized (scaled) the model predictions of TB incidence based on the pre-intervention local equilibrium and only considered active cases of the population (e.g., **Figure 2C&D**). The model dynamics proceeds differently in three separate time periods:

**Figure 2:**
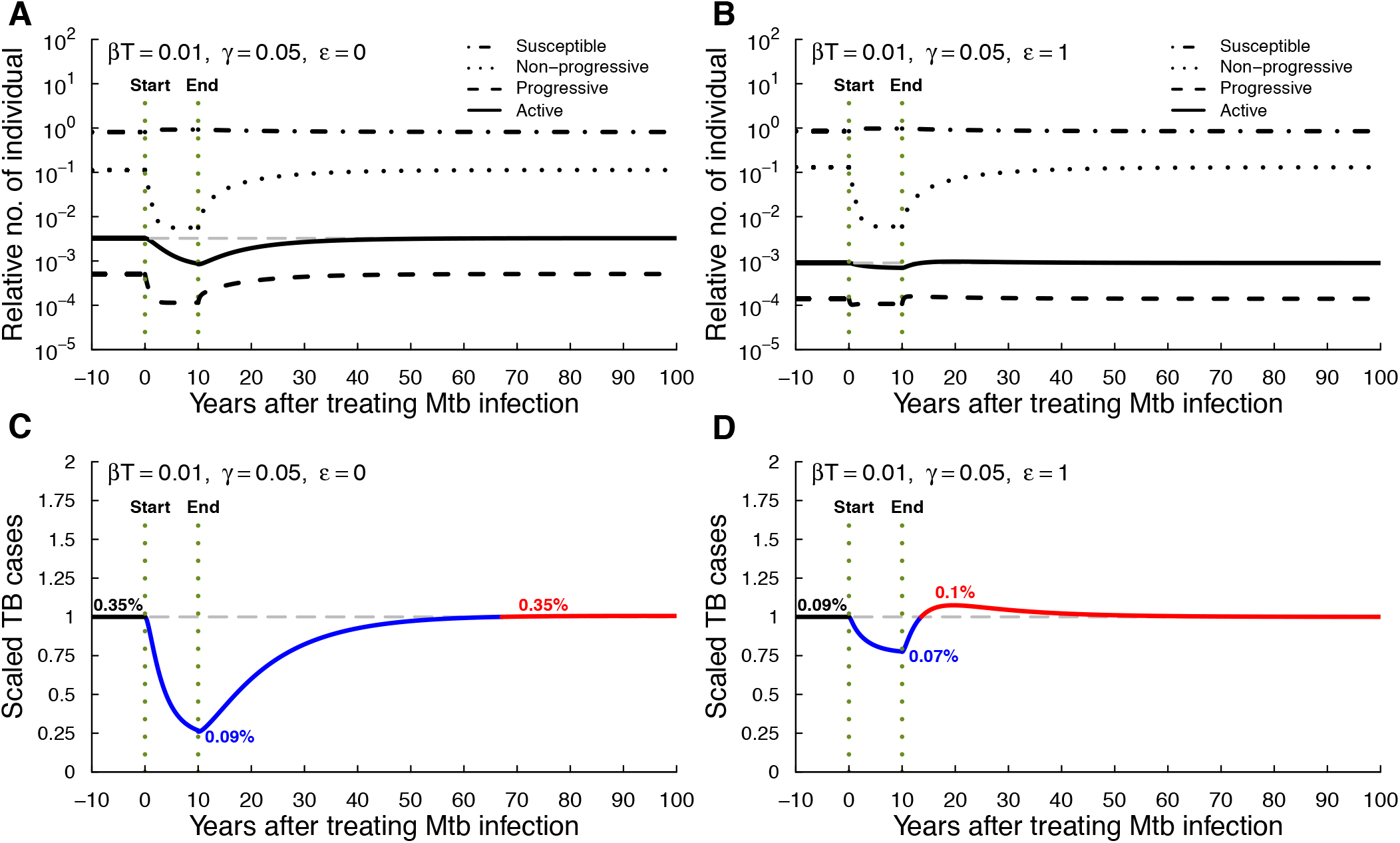
Stopping a program of community-wide preventive treatment of Mtb infection may result in increase in TB cases above pre-treatment program levels. We simulated TB dynamics (see **eqns. (1)–(4)**) in low TB prevalence settings, i.e., by assuming low rates of Mtb infection (*βT*) and recovery (*γ*, see **Supplemental Table 1**) for scenarios when Mtb infection does not protect against reinfection (*ϵ* = 0, **A&C**) or when Mtb infection is fully protective against reinfection (*ϵ* = 1, **B&D**). We show the dynamics of different sub-populations (**A&B**) or dynamics of TB cases (**C&D**), scaled to the pre-treatment steady state levels, prior to treatment program (years -10 to 0), during preventive treatment of asymptomatic individuals (years 0-10), or after the end of the treatment program (years 10-100). In panels (**C&D**) numbers indicate TB incidence in the population, and line colors indicate benefits (**blue**) or shortcomings (**red**) of the treatment program over 100 years. Vertical dashed lines indicate the times of start and end of the treatment program. We found similar results in setting of high transmission and recovery rates (**Supplemental Figure S3**) and in the model assuming *n >* 1 stages of progression towards active disease (**Supplemental Figures S1–S5**).

i. Before the treatment phase (years -10–0): During this period, the model is in a stable local equilibrium, where the proportion of susceptible individuals, MTBI cases, and active TB patients does not change. This timeline is used as a reference or baseline for assessing the impact of the intervention. It is interesting to note that the proportion of individuals in the progressive state is the lowest in frequency, even lower than the frequency of individuals with active disease (**Figure 2**A&B). This is because in the model progression to active disease is relatively fast (1-2 years) as compared to individuals with active disease who may live for several years in the absence of treatment (**Table 1**).
ii. During preventive treatment program (years 0–10): After the initiation of preventive treatment program, the frequency of MTBI^+^ individuals began to decline rapidly. In addition, the incidence of active TB also began to decline, as progression to active disease was broken by the preventive treatment. TB incidence fell below the previous baseline within a few years of the intervention, reflecting the immediate success of the preventive treatment. In settings of low Mtb transmission, the proportion of individuals with active TB disease becomes 0.09% for *ϵ* = 0 and 0.07% for *ϵ* = 1 (**Figure 2C&D**). Towards the end of this period we observed a gradual decrease in the population of MTBI^+^ individuals in a non-progressive state and a gradual increase in the number of susceptible individuals. This is due to natural demographic changes and the effect of successful treatment, as MTBI-induced immunity is lost as a result of treatment (**Figure 2B**).
iii. After discontinuation of preventive treatment program (years 10–100): After the TPT program was stopped, the model entered a new dynamic phase with predictions being strongly dependent on the degree of protection provided by MTBI (*ϵ*). If MTBI provides no protection against reinfection (*ϵ* = 0), stopping preventive treatment program will result in slow increase in TB cases that would reach pre-program levels in 60-70 years (**Figure 2C**). Interestingly, in the case high Mtb transmission, peak TB incidence slightly exceeds the pre-program levels (**Supplemental Figure S3C**). In contrast, when MTBI-induced protection is high (*ϵ* = 1), stopping the program results in rapid increase in TB incidence that exceeds pre-program levels less than 10 years after the program’s end (TB incidence is 0.1%, **Figure 2D**). In high transmission settings, rebound in TB cases approaches levels observed prior to program implementation (**Supplemental Figure S3D**).

In this model the dynamics of rebound of TB incidence in the population is driven by the changes in the number of susceptible individuals. In the case when MTBI does not provide protection against reinfection (*ϵ* = 0), preventive treatment program increases the number of susceptible individuals because fewer individuals progress to active disease. In the case when MTBI provides protection full against reinfection (*ϵ* = 1), preventive treatment program by treating individuals in non-progressive state making them susceptible to reinfection.

To confirm robustness of our predictions we performed similar simulations with our general model that includes *n* stages for individuals in progressive or non-progressive states (**Supplemental Figure S1** and **eqns. (S.1)–(S.6)**). Importantly, this slightly more complex model also predicted initial decline of TB incidence during the TPT program and slow or rapid rebound in TB cases after the program end (**Supplemental Figures S4 and S5**). Interestingly, this version of the model predicted higher than pre-program TB incidence in settings of high Mtb transmission and lack of MTBI-induced protection (**Supplemental Figure S4**C) illustrating the predicting impact of TPT program on TB incidence depends on modeling details.

### Increase in TB cases after preventive treatment program stop depends on the degree of protection provided by MTBI

We next extended our analyses to evaluate impact of preventive treatment program in the population with variable degree of MTBI-derived protection degree (*ϵ*) both in high (**Figure 3A**) and low (**Figure 3B**) Mtb transmission settings. We systematically varied the level of immunity provided by MTBI from *ϵ* = 0 (no protection) to *ϵ* = 1 (full protection). To characterize the impact of the preventive treatment program on TB incidence we calculated two characteristics of changes of TB incidence curve over time: an area over the curve (**AOC**) and an area under the curve (**AUC**). AOC measures impact of the program on decrease of the TB incidence and was calculated as an area under the TB incidence curve from year 0 until the curve returns to the pre-program levels. AUC measures increase in TB incidence after the treatment program end that are above the pre-program levels (**Figure 3**).

**Figure 3:**
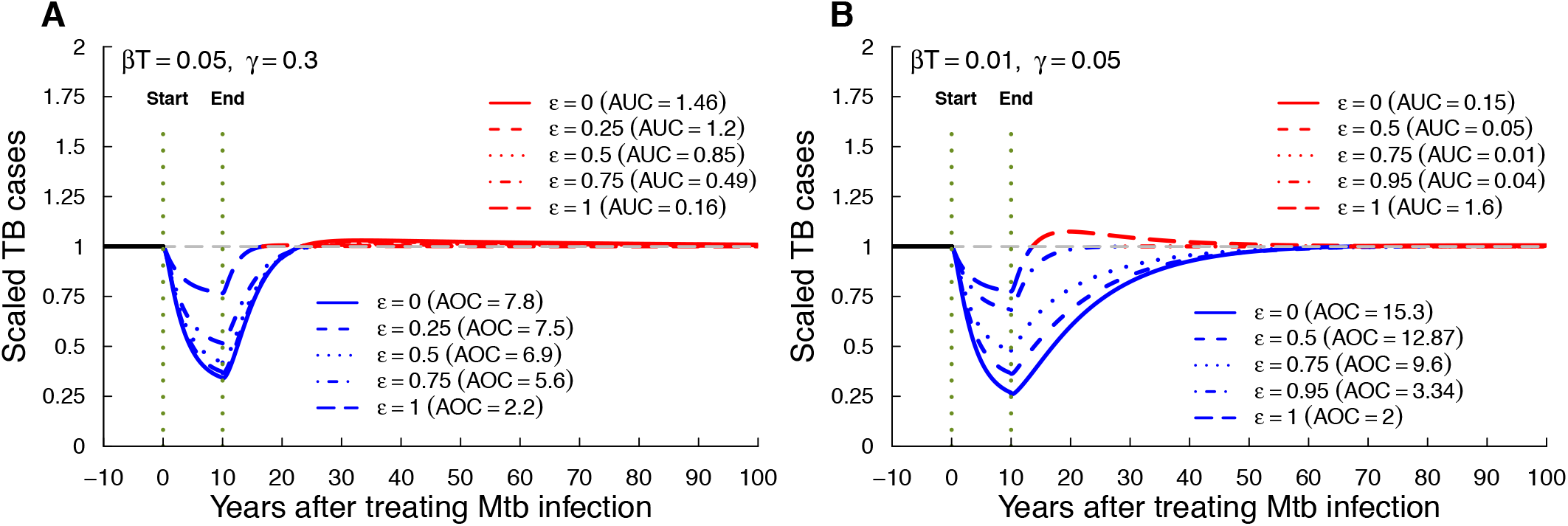
Rebound after stopping community-wide MTBI treatment program may occur with or without MTBI-induced protection. We simulated TB dynamics in high (**A**) or low (**B**) initial TB prevalence settings with varying the degree of protection *ϵ* afforded by MTBI (see **eqns. (1)–(4)** and **Supplemental Table 1**). We show changes in percent of TB cases scaled to its value prior to implementation of the treatment program (see **Figure 2** for detail). In panels, **AOC** is the area over the curve, representing the total (scaled) number of TB cases reduced due to preventive treatment program, and **AUC** is the area under the curve, representing the total (scaled) number of TB cases arising after stopping the treatment program. We found similar results in a model with *n* > 1 stages of progression towards active disease (**Supplemental Figure S6**).

Importantly, the degree of MTBI-induced protection had a large impact on dynamics of TB incidence during and after the program. The initial reduction in TB cases during treatment program was largest if MTBI does not protect against reinfection; for example, in high transmission setting

AOC values varied between 7.8 (*ϵ* = 0) to 2.2 (*ϵ* = 1), and in low transmission setting, AOC gradually changed from 15.3 to 2 for *ϵ* changing from 0 to 1, respectively (**Figure 3**). In contrast, increase in TB cases after the program end above the pre-program levels was dependent on both Mtb transmission rate and degree of MTBI-induced protection; in particular, at low Mtb transmission setting and high MTBI-induced protection, the increase in TB incidence after program end (AUC) nearly matched the reduction in TB cases during the program (AOC, **Figure 3**B). However, in high transmission setting the largest increase in TB incidence after program end was when MTBI-induced protection was small (**Figure 3**A). Importantly, large increase in TB incidence after program stop was only achieved at very high degree of MTBI-induced protection (*ϵ ≈* 1) and even at slightly lower values of *ϵ* (e.g., *ϵ* = 0.95), increase in TB incidence post program was small (**Figure 3**B). We reached similar conclusions when simulating Mtb dynamics in accord with the general *n*-stage model (**Supplemental Figure S6**).

### Infection rate (*βT*), recovery rate (*γ*) and rate of reversion to the non-progressive state (*λ*) impact differently benefits of the MTBI treatment program

Our previous analyses indicate that benefits of the community-wide preventive treatment program depend on Mtb transmission rate and on the degree of MTBI-induced protection (**Figure 3**). To further investigate the impact of variability in other parameters on TB incidence dynamics we randomly drew values for the infection rate *βT*, recovery rate (*γ*) and rate of reversion to the non-progressive state (*λ*) from a normal distribution, while keeping the remaining parameters fixed (see Materials and methods for detail), and simulated model dynamics. To quantify impact of the treatment program on TB incidence we calculate three summary metrics based on the time-dependent change in active TB cases such as AOC, AUC, and the net value Δ = AUC-AOC (**Figure 3**). Negative values of Δ indicate the overall benefits of the preventive treatment programs (decline in TB cases is larger than increase after the program end), while positive Δ indicates net detrimental effects of the TPT program.

In these simulations we also found that the overall benefit of the preventive treatment program depends on the degree of MTBI-induced protection (**Figure 4**). When MTBI is not protective (*ϵ* = 0), reduction in TB cases during the treatment program is substantial reaching 200% in low transmission settings (blue dashed lines in **Figure 4**A&C). While there may be substantial rebound in TB incidence after the program end, it typically did not exceed the early benefits of the program leading to relatively high net benefits (black lines in **Figure 4**A&C). In contrast, when MTBI is fully protective (*ϵ* = 1), reduction in TB cases due to the program is relatively small but predicted increase in TB incidence after program end may be substantial (**Figure 4**B&D). In fact, in settings of low Mtb transmission, the net effect of the treatment program may be positive with substantial increase in TB incidence 90 years after the program end (**Figure 4**D). Our results were similar for the general *n* stage model of Mtb dynamics (**Supplemental Figure S7**).

**Figure 4:**
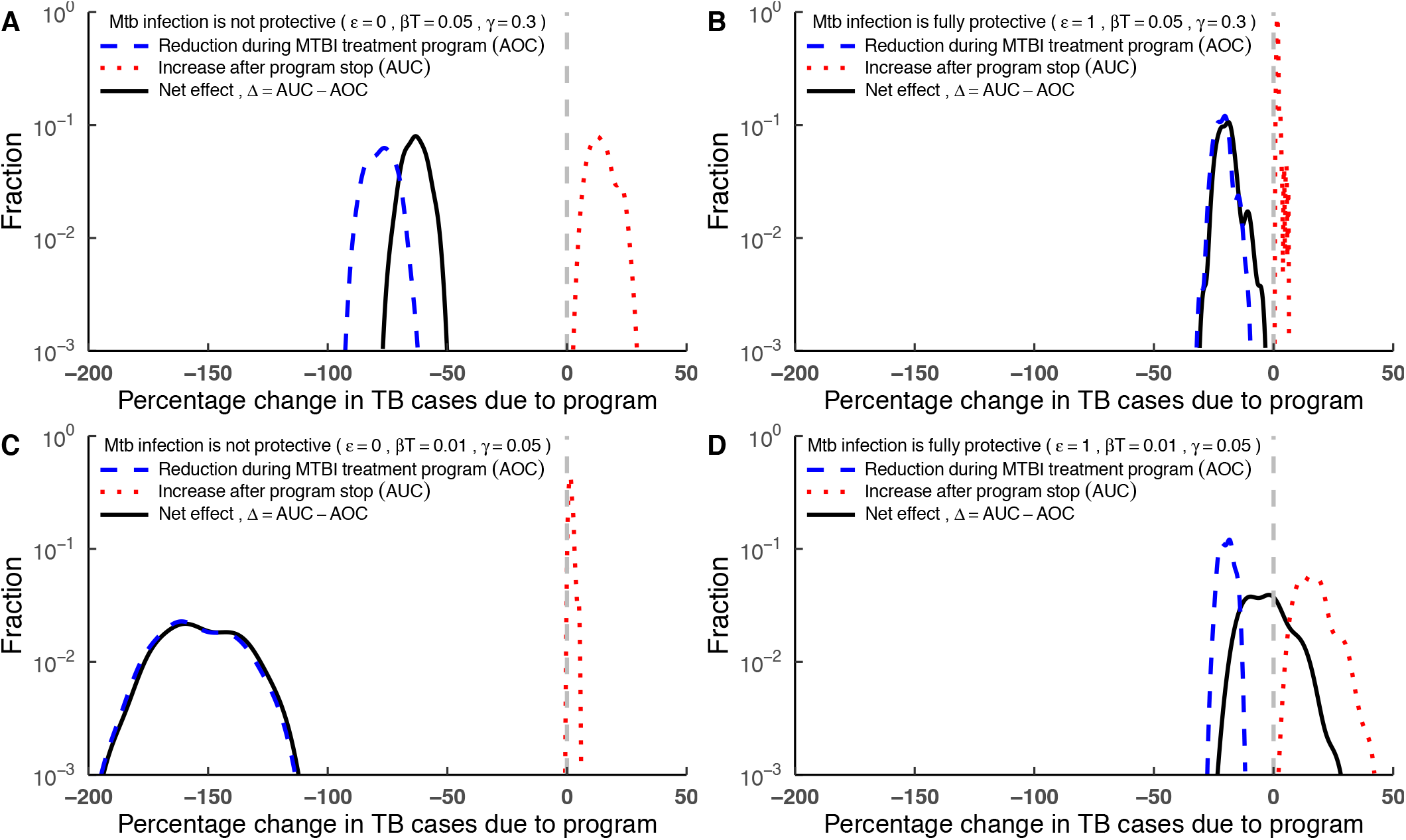
Benefits of MTBI treatment program depend on TB prevalence and immunological details. We simulated infection dynamics using **eqns. (1)–(4)** with randomly (normal distribution) sampled parameters with averages given in **Supplemental Table 1** and *σ* = 0.2 in 100 simulations when MTBI is non protective (*ϵ* = 0, **A&C**) or fully protective (*ϵ* = 1, **B&D**) and in high (**A**-**B**) or low (**C**-**D**) transmission and recovery rates. **AOC** and **AUC** represent area over the curve (reduction in TB cases due to TPT program) and area under the curve (increase in TB cases above the pre-treatment levels due to rebound), respectively, and Δ = AUC *−* AOC is a net benefit of the preventive treatment program. We calculated the percent change in TB cases (x axis values) by scaling values of AOC or AUC by 10 that is the best possible outcome of the treatment program that would reduce (scaled) TB incidence from one to zero for 10 years of the program (see **Figure 3** for detail). We found similar results in the model assuming that progression to active disease occurs via *n* > 1 stages (**Supplemental Figure S7**).

### Intuitive explanations of rebound in TB incidence after stopping community-wide preventive program

We found that under certain parameter settings stopping MTBI treatment program can lead to a higher TB incidence than that existed prior to implementing the program. This outcome can be explained intuitively by the following mechanisms that are dependent on the degree of MTBI-induced protection:

a. **Immunity loss in treated MTBI**^+^ **individuals (***ϵ ≈* 1): When *ϵ ≈* 1, MTBI provides significant protection against reinfection and progression to active disease. Treatment of MTBI will inadvertently eliminate this natural protection and will increase the pool of individuals fully susceptible to reinfection. As the preventive treatment program is stopped such individuals would progress to TB rapidly (**Figure 5A**).
b. **Population structural changes and reduction of TB mortality (***ϵ ≈* 0): In the model, MTBI treatment significantly reduce the overall mortality due to TB in the population. While this is positive in the short term, in the long term it increases the number of susceptible individuals in the population. After stopping the TPT program, these susceptible individuals become vulnerable to reinfection, resulting in increase in TB incidence (**Figure 5B**).

**Figure 5:**
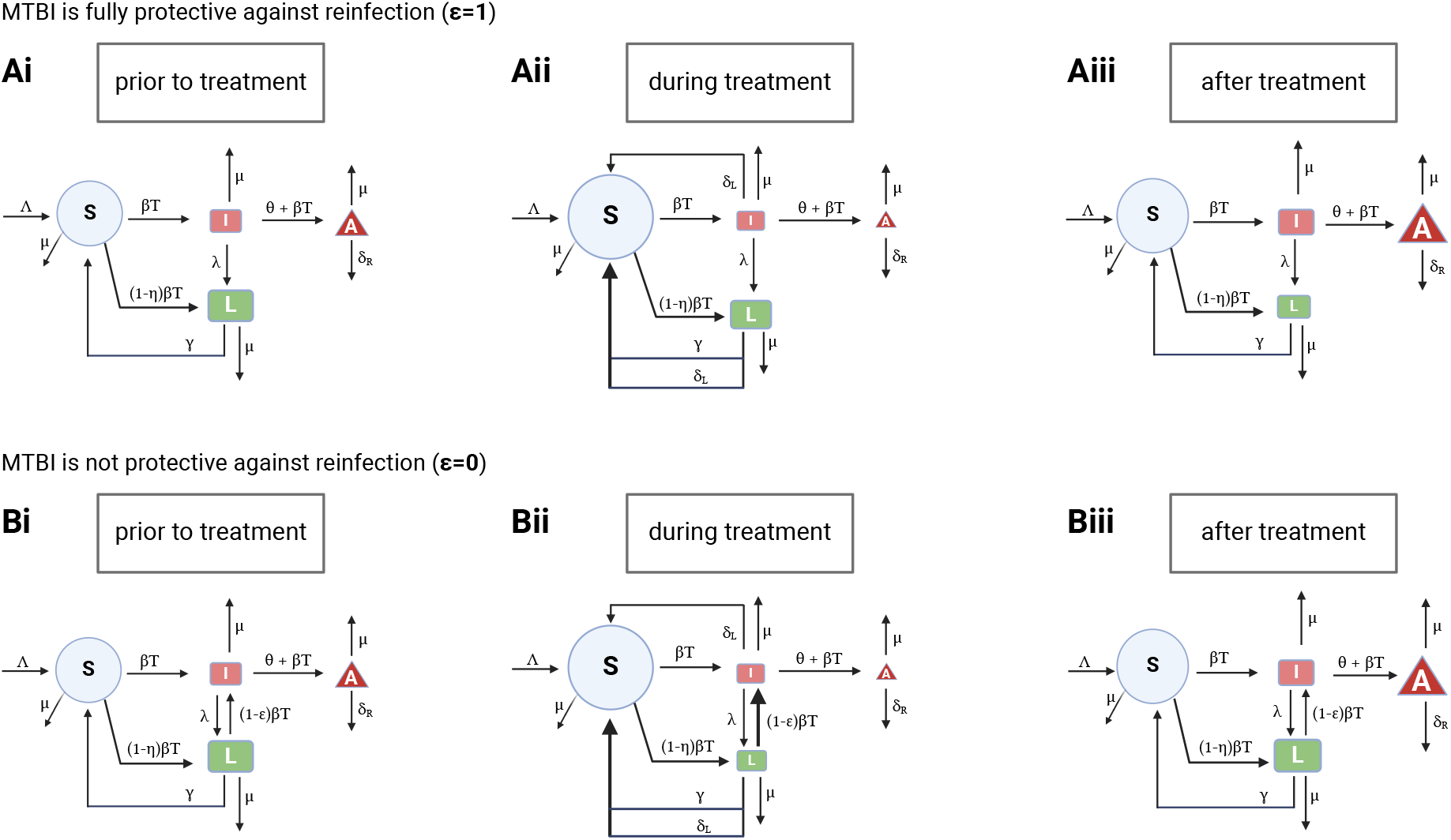
Schematic illustrating intuitive explanation of the large increase in TB cases after stopping preventive treatment of MTBI. We show three outlines of the community structure before MTBI treatment program (**Ai & Bi**), during MTBI treatment program (**Aii & Bii**) and after MTBI treatment program is stopped (**Aiii & Biii**) when MTBI fully protects against reinfection (**A**, *ϵ* = 1, low infection and recovery rates) and when MTBI is not protective (**B**, *ϵ* = 0, high infection and recovery rates, see **Supplemental Table 1**). The size of the circles/boxes indicate the relative (not to scale) proportions of different subpopulations prior, during, and after treatment program, and thickness of the arrows also indicates their relative values. **A**: In low TB prevalence settings when non-progressive individuals are fully protected against reinfection (*ϵ* = 1), treatment of asymptomatic individuals results in increase in the number of individuals susceptible to infection (**Aii**). These individuals become infected and progress to active disease after treatment program stops (**Aiii**). **B**: In high TB prevalence settings when non-progressive individuals can be easily reinfected with Mtb (and thus transition to the progressive state), treatment of asymptomatic individuals results in increase in the number of individuals susceptible to infection and in individuals in non-progressive state; these changes are also driven by the reduction in overall mortality due to reduced numbers of active cases (**Bii**). Stopping the treatment program results in increase in active cases because both susceptible individuals and individuals in non-progressive state become reinfected and progress to active disease (**Biii**).

## Discussion

In this paper we investigated potential benefits and shortcomings of a community-wide treatment program to prevent TB development in individuals with evidence of past or present MTBI. Our analysis of the data from a large cohort of nursing students in the pre-antibiotic era suggests that TST^+^ students developed active disease more slowly than TST^*−*^ students; we estimated that TST positivity provided *ϵ ≈* 65% reduction in the rate of progression to active disease (**Supplemental Figure S2**C). While we did not find a statistically significant difference in the cumulative proportion of TST^*−*^ or TST^+^ students who developed TB (40/322 vs. 31/343, 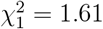, *p* = 0.21), other studies have found such differences^43,49,50^, and meta-analysis of multiple cohort studies suggests that MTBI may provide 80% protection against active disease upon re-infection^52^.

We developed a novel mathematical model of TB epidemiology by dividing MTBI^+^ individuals into those on a path of progression towards active disease and those who are in a non-progressive state and may benefit from MTBI-induced protection when re-exposed to Mtb (**Figure 1** and **eqns. (1)–(4)**). In the model, a program where all MTBI^+^ individuals receive preventive treatment (and all are efficiently identified and compliant with treatment) is highly beneficial at reducing TB incidence (e.g., **Figures 2 and 3**); however, stopping such a program if the overall Mtb transmission is not reduced will result in rebound of TB incidence that may exceed pre-program levels (**Figure 3**). Both Mtb transmission rate and the degree of protection provided by MTBI play a key role in the magnitude of the rebound (**Figure 4**) although the overall mechanism is somewhat similar (increase in the number of susceptible individuals, **Figure 5**).

Our approach to model Mtb dynamics and disease progression in a community builds upon many previous studies of TB epidemiology ^6,23,24,58–62^ and extends this previous work in several important directions. First, in contrast with previous models we do not assume that Mtb-infected individuals always ultimately progress to active disease. Individuals who become Mtb infected but do not progress to active disease then can be protected against re-infection and switch to a progressive state. Cohort studies in which even individuals with evidence of MTBI (e.g., TST^+^ individuals) progress to active disease in high Mtb exposure settings are consistent with this model assumption. Second, we assume that in the absence of reinfection, MTBI^+^ individuals may eventually control (and perhaps eliminate) the infection and thus revert to the non-immune/susceptible state. While the ability of TST^+^ individuals to lose their Mtb reactivity over time has been recognized for a long time^43^, mathematical models have rarely included this mechanism^24,66^. Third and finally, even though previous models have included treatment of individuals with MTBI^6^, these models did not consider loss of immunity against reinfection following the treatment.

While there have been calls for wider implementation of preventive treatment in different communities^6,8,9,82,83^, the overall effectiveness of community-wide preventive treatment programs in reducing TB incidence remains debated. Early clinical trials of 12-month preventive treatment with INH showed lasting benefits in multiple settings^34^; however, more recent similar clinical trials in South Africa including preventive treatment of gold miners or individuals with a gene signature-based high risk of TB progression did not show long-term benefits^35,36^. The latter findings are not fully consistent with predictions of our perhaps idealized model that showed reduction in active disease early during preventive treatment period (e.g., **Figure 2**). Rapid rise in TB incidence in individuals undergoing preventive treatment after the treatment was stopped in clinical trials^36,84^ is somewhat consistent with our modeling predictions in scenarios of high degree of protection provided by MTBI and its loss upon preventive treatment.

The degree of protection provide by MTBI has been debated^29,85^. One of the earliest observations that individuals with MTBI (detected as TST positivity) have a lower likelihood of developing TB comes from Prophit survey that followed 10,000 individuals for 10 years from 1935 to 1944 (ref ^43^). It is interesting to note that while Daniels *et al*. ^43^ acknowledged the possibility that MTBI may be protective against Mtb re-infection, the authors provided rigorous justification that MTBI^+^ individuals may be simply resistant to TB, in part, due to selection via early mortality of TB-susceptible individuals. Given reduction in childhood mortality due to TB because of BCG vaccination^86^, detection of MTBI-induced protection in more recent studies is more likely because of MTBI-induced immunity and not due to intrinsic resistance to TB^52^.

Our work has several limitations. The updated WHO guidelines recommend treatment for individuals who are at the higher risk of progression to TB upon exposure to Mtb, e.g., contact of TB cases especially children, people living with HIV or immunocompromised individuals ^1^. Our mathematical models currently ignore such specific subgroups. We assume that the force of infection *βT* is not influenced by the preventive treatment program, i.e., is independent of the number of individuals with active disease in the community (see **eqns. (1)–(4)**). Such an assumption may overestimate the increase in TB incidence after the program is stopped and may be only valid if the members of the community remain exposed to Mtb from other sources. While most of our model parameters are taken from previously published studies (see Materials and methods), their values may not fully represent a given community, and are likely to be unique for different communities/countries. Impact of treating MTBI^+^ individuals on TB prevalence is likely to depend on these specific parameters. Protection afforded by MTBI against reinfection may be individual and time-dependent. Furthermore, how the protection afforded by MTBI changes in individuals under preventive treatment remains poorly understood. In our model we assume that immunity declines relatively quickly after treatment. However, this simplification may not fully reflect the dynamics of real immune responses and may affect our model’s estimates of long-term changes in TB incidence.

Our work opens avenues for future research. Extending mathematical models to consider different sub-populations in a community (e.g., children, people living with HIV) including by using agent-based models (**ABM**) is likely help quantify the role of population heterogeneity on TB incidence following implementation of community-wide preventive treatment program. Better understanding how MTBI provides protection against Mtb reinfection and how preventive treatment may impact MTBI-induced protective immunity in humans is desperately needed, including in setting of aging, HIV infection, and other comorbidities. Extending the model to allow transmission to be maintained by the prevalence of individuals with active disease (i.e., using *βA* instead of *βT* in **eqns. (1)–(3)**) and including asymptomatic infectious individuals would be important to more rigorously quantify impact of the community-wide preventive treatment programs on TB prevalence.

Taken together, our results raise a concern with predicting the long-term impact of community-wide preventive treatment on TB prevalence. Although preventive treatment can reduce the burden of disease in the short term, if it is discontinued without monitoring and complementary interventions (such as vaccination or active disease detection), it may unintentionally worsen the epidemic situation. Our results also demonstrate how the interplay between infection intensity, recovery rate, and MTBI-induced immunity profoundly influences the success or failure of TB control strategies. This model-based analysis is also consistent with previous studies that have warned of the risk of abrupt cessation of TB control programs, especially in areas with high transmission rates and partial immunity from previous infection^87^.

## Data Availability

All data produced in the present study are available upon reasonable request to the authors.

https://github.com/stbatabyal/latency_protection/tree/main

## Abbreviations

Mtb: Mycobacterium tuberculosis
TB: tuberculosis
MTBI: Mtb infection
TPT: TB preventive treatment
WHO: World Health Organization
TST: tuberculin skin test
IGRA: interferon gamma release assay
AOC: area over the curve
AUC: area under the curve
LRT: likelihood ratio test
CoMtb: contained/concomitant Mtb

## Data and code sources

The data from the paper along with the codes are available on github: https://github.com/stbatabyal/latency_protection/tree/main. Analyses have been done in R (4.1.2) and Mathematica (ver 12).

## Author contributions

SB primarily did the modeling, computational simulations and wrote the first draft of the paper. KU contributed to the initial idea of the project. VVG performed analyses of experimental data. All authors read, edited, and agreed on the final version of the paper.

## Acknowledgments

We would like to thank Burroughs Wellcome Fund 2018 Collaborative Research Travel Grant that supported sabbatical of one of us (VVG) that led to this work. The authors thank Prof. M.A. Behr for his valuable comments that help to improve the abstract of this manuscript. Nasif Mohd from Fulton County Schools Internship program and Aryan Kumar from UTK contributed with preliminary works, and Dr. K. Avilov provided feedback on earlier versions of the paper.

## Supplemental Information

### General mathematical models of Mtb epidemiology

We extended our basic, one stage mathematical model, describing Mtb dynamics in a community to allow for *n* stages in both progressive (*I*_*j*_) and non-progressive (*L*_*j*_) states. The mathematical model is written as follows (**Supplemental Figure S1**):

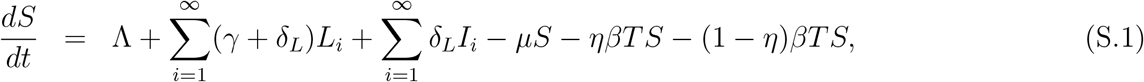

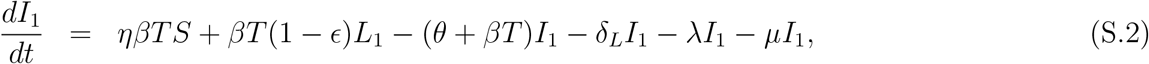

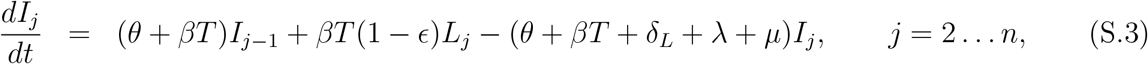

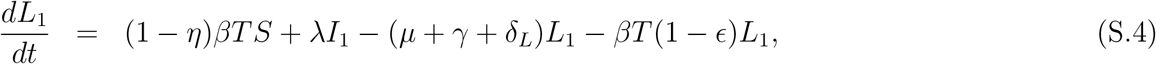

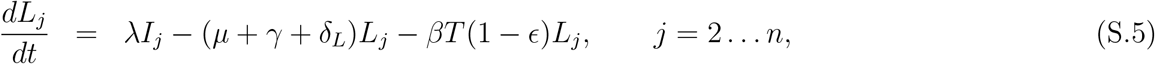

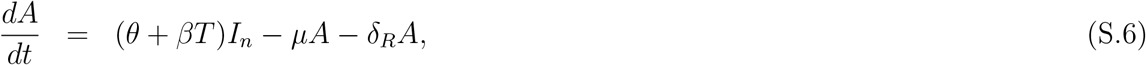

where *I*_*j*_ and *L*_*j*_ are the number of individuals in the *j*^th^ stage of the progressive and non-progressive states, respectively, and other notations are identical to **eqns. (1)–(4)**. The total number of individuals in the progressive and non-progressive states then are 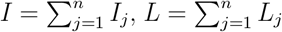 and *j* = 1 … *n*. Note that in this model to allow for 0.5-2 years for progression from infection to active disease upon primary exposure the progression rate *θ* for *n*-stage model must be scaled appropriately (typically, the rate is *n* times higher in the *n*-stage model as compared to the one-stage model, **Supplemental Table 1**).

**Supplemental Figure S1:**
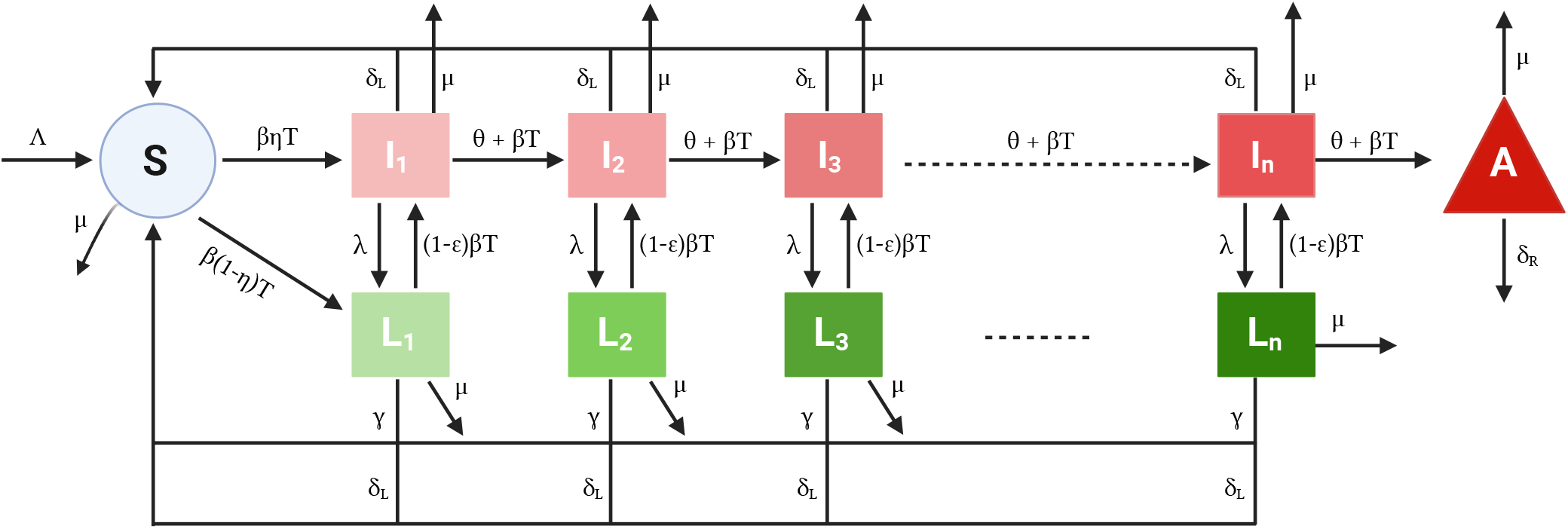
Schematic diagram of Mtb infection dynamics in a human population. We illustrate the *n*-compartment model with progressive and non-progressive states. Details of the schematic has been discussed in **Figure 1** and model equations are shown in **eqns. (S.1)–(S.6)**.

**Supplemental Figure S2:**
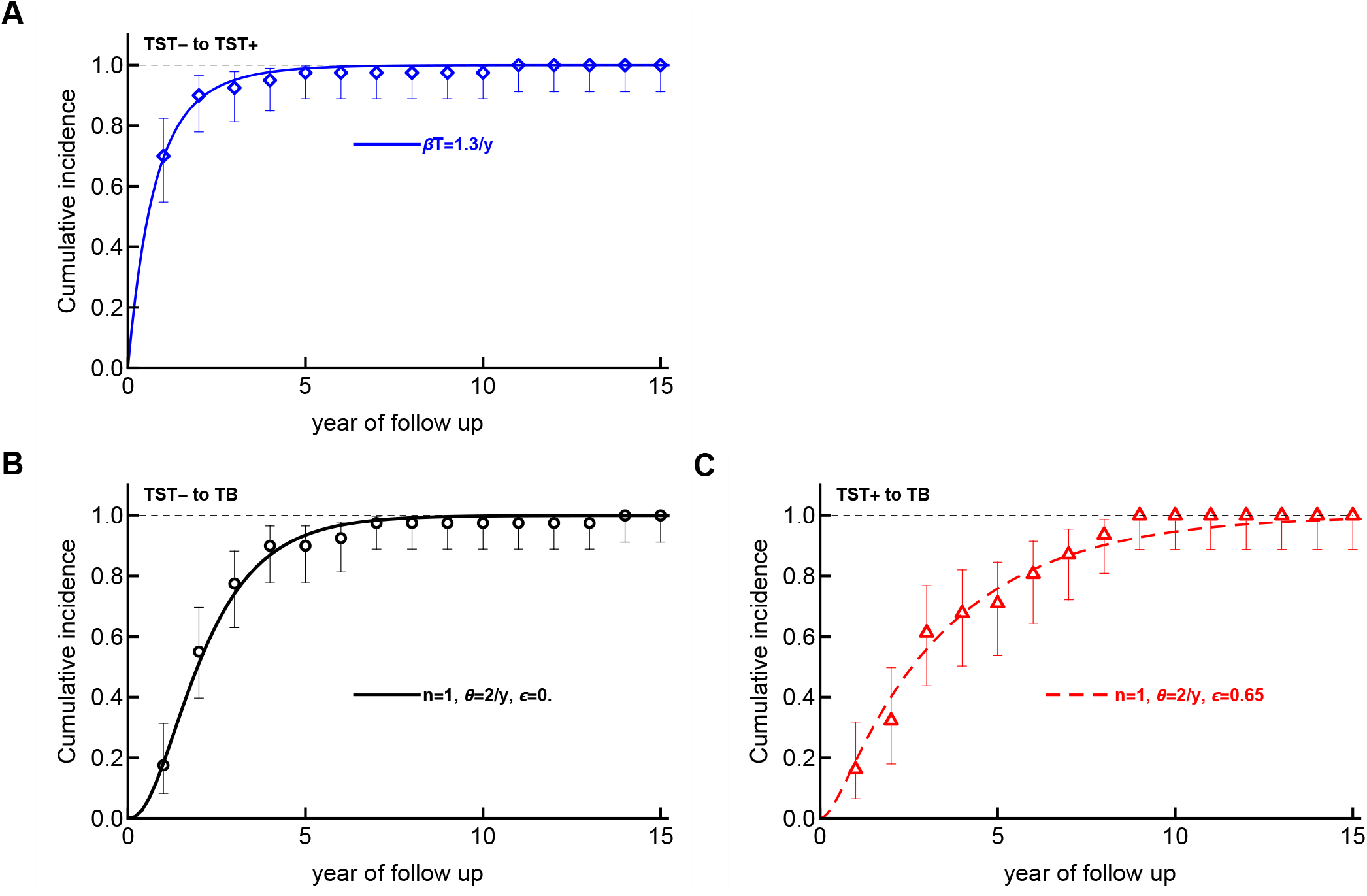
One stage epidemiological model for Mtb dynamics explains the data on TST conversion and progression to TB of a cohort of nursing students. We searched for parameter combinations of the basic epidemiological model (**eqns. (1)–(4)**) that explain the data for TST^*−*^individuals to become TST^+^ (**A**), and for TST^*−*^(**B**) and TST^+^ (**C**) individuals to progress to TB. In the model the *S/*(*S* + *I* + *L* + *A*) represents TST^*−*^individuals, (*I* + *L*)*/*(*S* + *I* + *L* + *A*) are TST^+^ individuals (with no TB). Parameters providing the best match of the model predictions (shown by lines) to data (shown by markers) are: *βT* = 1.3*/*y, *θ* = 2*/*y, *λ* = 0.5*/*y, *γ* = 0.2*/*y, and *ϵ* = 0.65 in **C**.

**Supplemental Figure S3:**
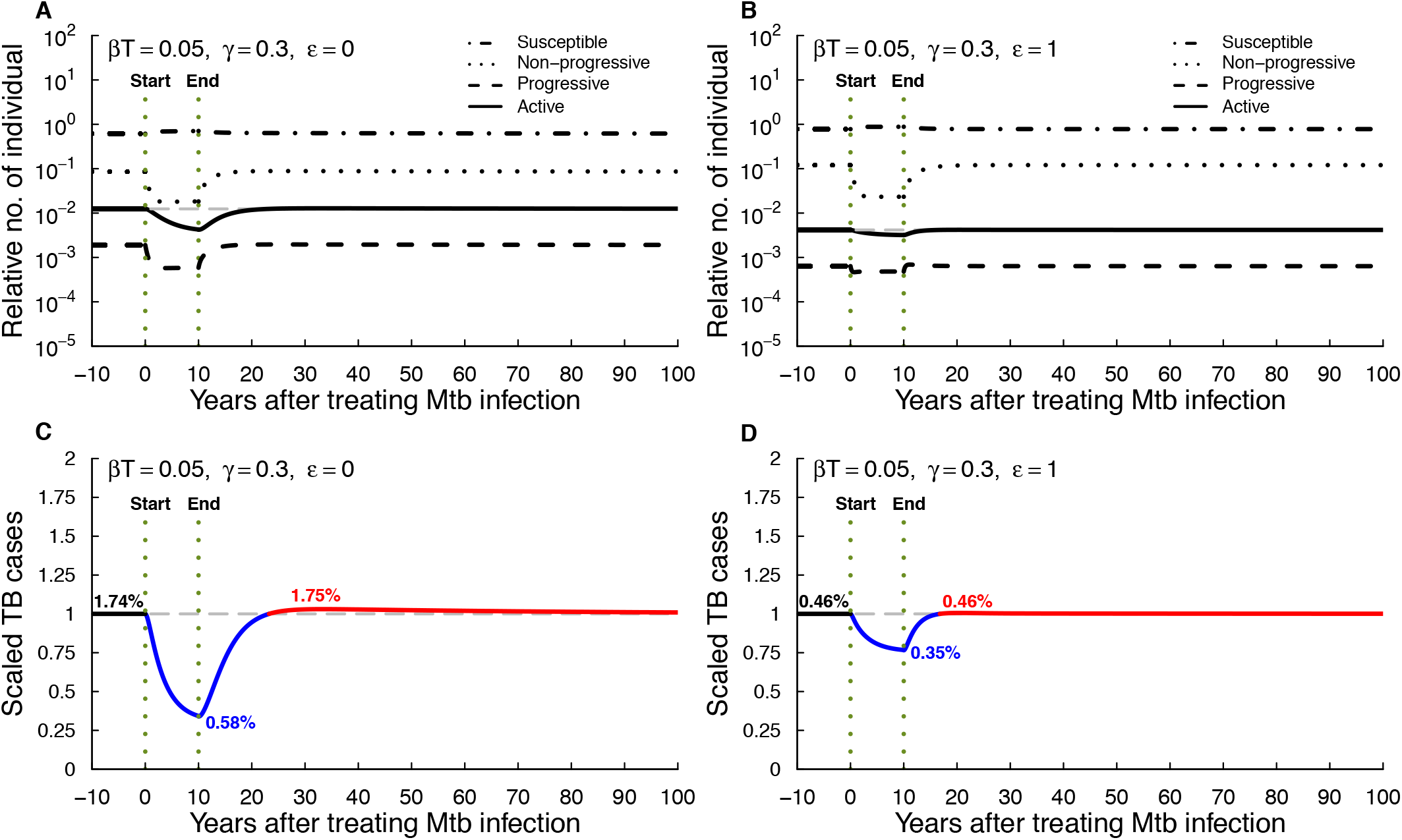
Stopping community-wide treatment of Mtb infection may results in increase in TB cases later. We simulated dynamics using **eqns. (1)–(4)** (model with one stage) assuming high rates of Mtb infection and recovery (**Table 1**) for scenarios when Mtb infection does not protect against reinfection (*ϵ* = 0, **A&C**) or when Mtb infection is fully protective against reinfection (*ϵ* = 1, **B&D**). Here *βT* = 0.05*/*year and *γ* = 0.3*/*year and other key details are discussed in **Figure 2**.

**Supplemental Table S1:** Parameters of the extended mathematical model assuming *n* = 5 stages for progressive and non-progressive states. The parameters are similar to those given for a one stage model (**Table 1**) except of the infection rate *βT*, recovery rate *γ*, and transition rate *θ*. Model is given in **eqns. (S.1)–(S.6)** (**Supplemental Figure S1**) assuming *n* = 5. Example simulations are shown in **Supplemental Figures S4–S7**.

| Representation of parameters | Parameters | High infection rate $\beta T$<br>High recovery rate $\gamma$ | | Low infection rate $\beta T$<br>Low recovery rate $\gamma$ | |
| --- | --- | --- | --- | --- | --- |
| Birth rate | $\Lambda, 1/y$ | 0.013 | 0.013 | 0.013 | 0.013 |
| Death rate | $\mu, 1/y$ | 0.013 | 0.013 | 0.013 | 0.013 |
| Fraction to progressive state | $\eta$ | 0.05 | 0.05 | 0.05 | 0.05 |
| Infection rate | $\beta T, 1/y$ | 0.15 | 0.15 | 0.01 | 0.01 |
| Recovery rate | $\gamma, 1/y$ | 0.9 | 0.9 | 0.05 | 0.05 |
| Progression rate to disease | $\theta, 1/y$ | 10 | 10 | 10 | 10 |
| Reversion to non-progressive state | $\lambda, 1/y$ | 1 | 1 | 1 | 1 |
| Rate of active TB to be removed | $\delta_R, 1/y$ | 0.3 | 0.3 | 0.3 | 0.3 |
| Rate of preventive treatment | $\delta_L, 1/y$ | 1.5 | 1.5 | 1.5 | 1.5 |
| Infection-induced protection | $\epsilon$ | 0 | 1 | 0 | 1 |
| **% TB** | - | **$\epsilon = 0$** | **$\epsilon = 1$** | **$\epsilon = 0$** | **$\epsilon = 1$** |
| Prior to treatment (Steady states) | - | 4.81% | 1.26% | 0.33% | 0.08% |
| During treatment (Nadir) | - | 1.69% | 0.76% | 0.07% | 0.05% |
| After treatment (Peak ) | - | 4.84% | 1.26% | 0.33% | 0.09% |

**Supplemental Figure S4:**
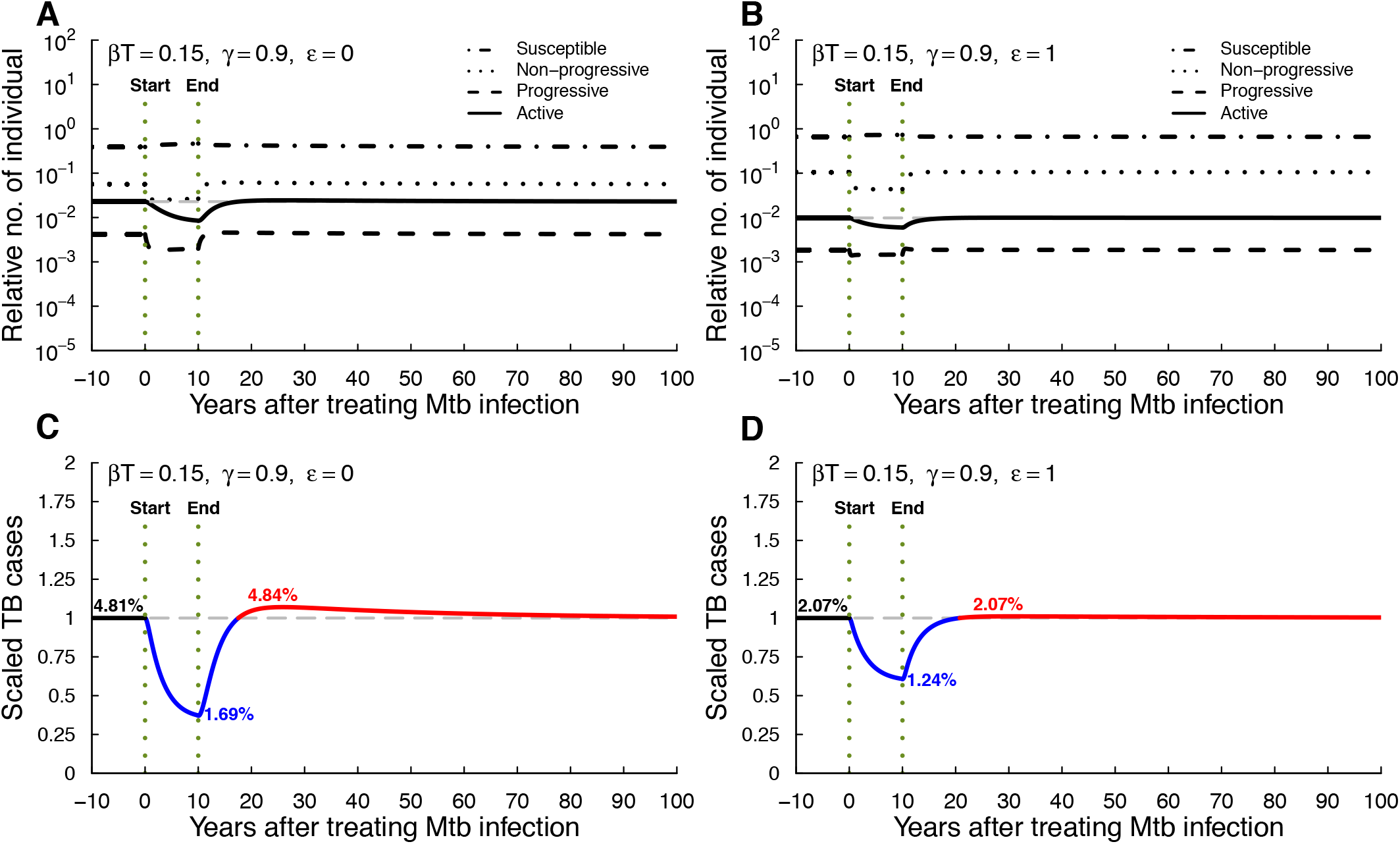
Stopping community-wide treatment of Mtb infection may results in increase in TB cases later. We simulated dynamics using a model with *n* = 5 stages for progressive and non-progressive states (see **eqns. (S.1)–(S.6)** and **Supplemental Figure S1**) with high rates of Mtb infection and high recovery rate (**Supplemental Table S1**) for scenarios when Mtb infection does not protect against reinfection (*ϵ* = 0, **A&C**) or when Mtb infection is fully protective against reinfection (*ϵ* = 1, **B&D**). Here *βT* = 0.15*/*year, *γ* = 0.9*/*year and other key details are discussed in **Figure 2**.

**Supplemental Figure S5:**
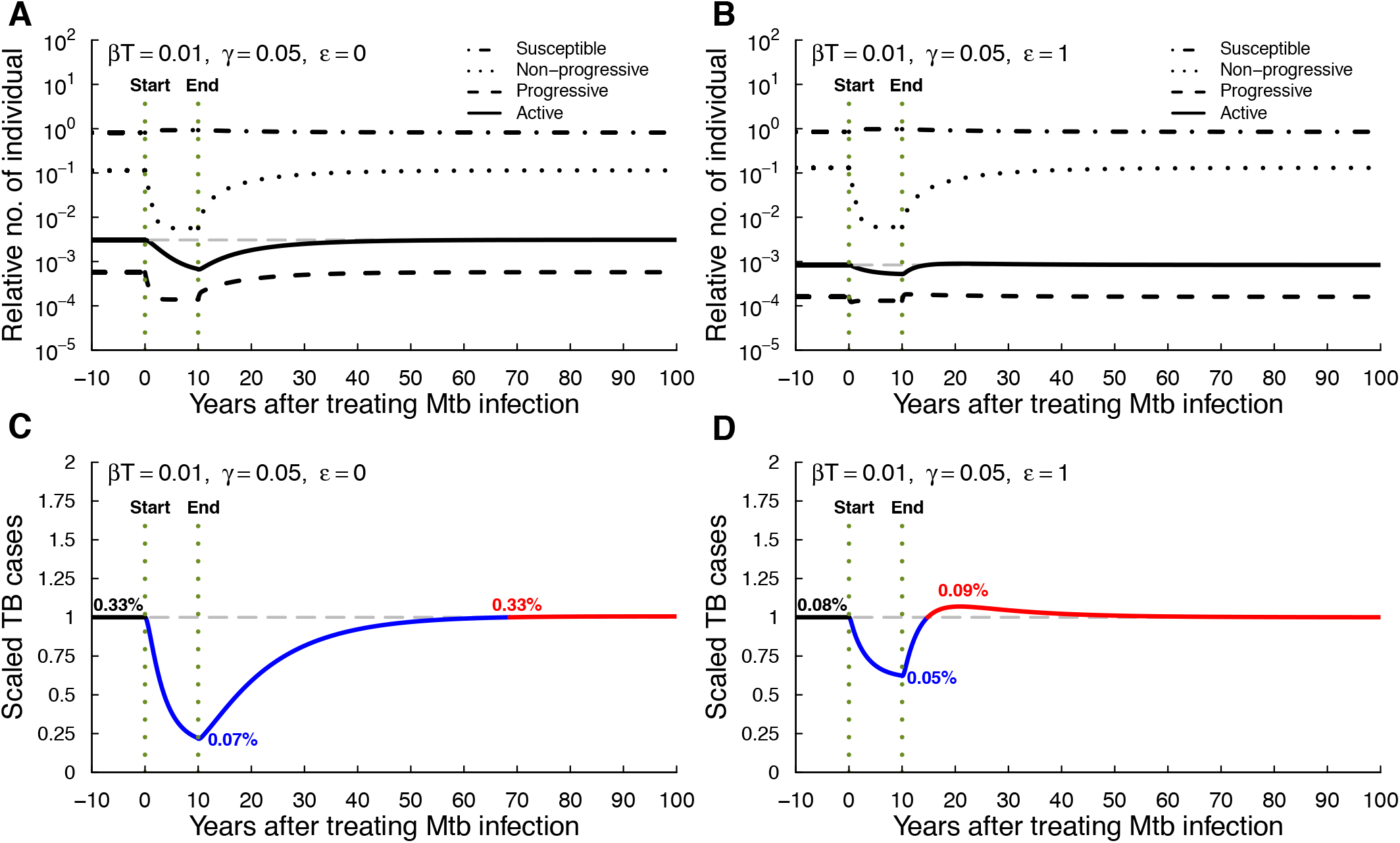
Stopping community-wide treatment of Mtb infection may results in increase in TB cases later. Similar simulations as in **Supplemental Figure S4** except with low rates of Mtb infection and low recovery rate (**Supplemental Table S1**) for scenarios when Mtb infection does not protect against reinfection (*ϵ* = 0, **A&C**) or when Mtb infection is fully protective against reinfection (*ϵ* = 1, **B&D**). Here *βT* = 0.01*/*year, *γ* = 0.05*/*year.

**Supplemental Figure S6:**
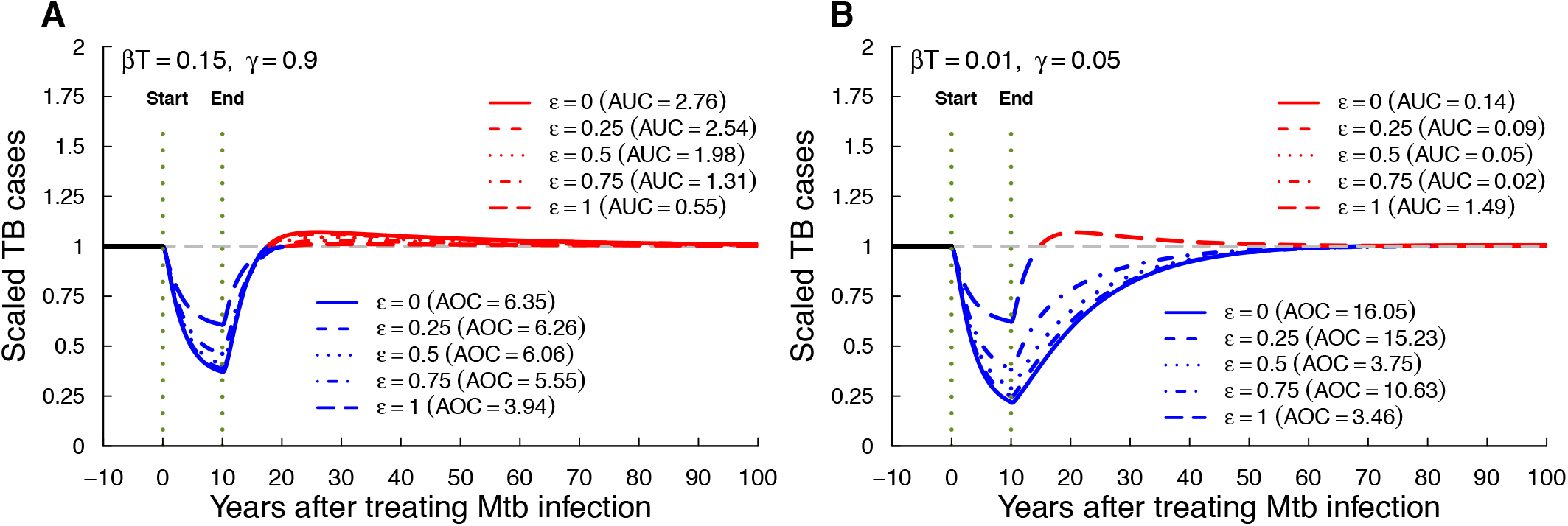
Rebound after stopping community-wide MTBI treatment program may occur with or without infection-induced protection. We simulated dynamics using a model with *n* = 5 stages for progressive and non-progressive states (see **eqns. (S.1)–(S.6)**) and calculated the kinetics of active cases over time (see **Figure 3** for detail). (**A**: high infection and recovery rates; **B**: low infection and recovery rates) and varied the degree of protection afforded by the MTBI (*ϵ*). Here *βT* = 0.15*/*year & *γ* = 0.9*/*year (A), *βT* = 0.01*/*year & *γ* = 0.05*/*year (B).

**Supplemental Figure S7:**
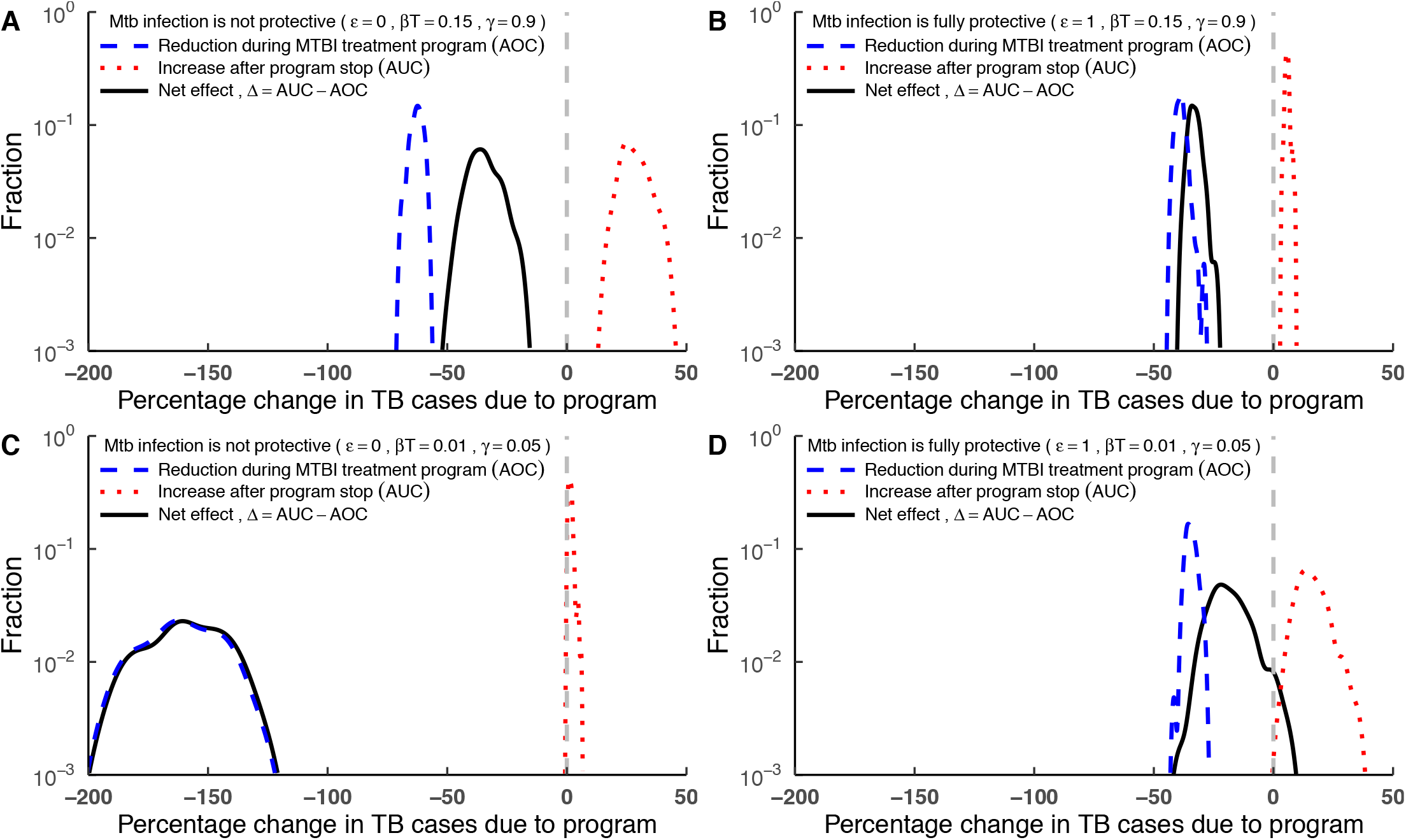
Benefits of MTBI treatment program depend on immunological details. Similar simulations to those in **Figure 4** but with a model considering *n* = 5 stages (see **eqns. (S.1)–(S.6)** and **Supplemental Figure S1**). We simulated infection dynamics with normally sampled parameters with averages given in **Supplemental Table S1** and *σ* = 0.2 in 100 simulations when MTBI is non protective (*ϵ* = 0) and fully protective (*ϵ* = 1) with high infection and recovery rates (*βT* = 0.15*/*year and *γ* = 0.9*/*year, **A&B**) or non protective (*ϵ* = 0) and fully protective (*ϵ* = 1) with low infection and recovery rates (*βT* = 0.01*/*year and *γ* = 0.05*/*year, **C&D**). Other key details are described in **Figure 4**.

